# SARS-CoV-2 Omicron XBB infections boost cross-variant neutralizing antibodies, potentially explaining the observed delay of the JN.1 wave in some Brazilian regions

**DOI:** 10.1101/2024.09.28.24314453

**Authors:** Luis Fernando Lopez Tort, Mia Ferreira de Araújo, Ighor Arantes, Jéssica SCC Martins, Marcelo Gomes, Felipe Cotrim de Carvalho, Walquiria Aparecida Ferreira de Almeida, Braulia Costa Caetano, Luciana R. Appolinario, Elisa Calvalcante Pereira, Jéssica Carvalho, Fábio Miyajima, Gabriel Luz Wallau, Felipe Gomes Naveca, Pedro Alves, Otávio Espíndola, Patricia Brasil, Paola Cristina Resende, Gonzalo Bello, Marilda Mendonça Siqueira

## Abstract

**Objectives:** The SARS-CoV-2 JN.1 lineage emerged in late 2023 and quickly replaced the XBB lineages, becoming the predominant Omicron variant worldwide in 2024. We estimate the epidemiological impact of this SARS-CoV-2 lineage replacement in Brazil and we further assessed the cross-reactive neutralizing antibody (NAb) responses in a cohort of convalescent Brazilian patients infected during 2023.

**Methods:** We analyzed the evolution of SARS-CoV-2 lineages and severe acute respiratory infection (SARI) cases in Brazil between July 2023 and March 2024. We evaluated the cross-reactive NAb responses to the JN.1 variant in a cohort of convalescent Brazilian patients, both before and after infection with XBB.1* lineages.

**Results:** JN.1 replaced XBB with similar temporal dynamics across all country regions, though its epidemiological impact varied between locations. The Southeastern, Southern, and Central-Western regions experienced a brief XBB wave around October 2023, shortly before the introduction of JN.1, without any immediate upsurge of SARI cases during viral lineage replacement. By contrast, the Northeastern and Northern regions did not experience an XBB wave in the latter half of 2023 and displayed a rapid surge in SARI cases driven by the emergence of the JN.1. We found that recent XBB infections in the Brazilian population significantly boosted cross-reactive NAb levels against JN.1.

**Conclusions:** The XBB wave observed in the second half of 2023 in some Brazilian states likely acted as a booster for population immunity, providing short-term protection against JN.1 infections and delaying the rise of SARI cases in certain regions of the country.

## Introduction

In response to the decline in Coronavirus disease 2019 (COVID-19) related hospitalizations and intensive care unit admissions, and the high rates of population immunity against the Severe Acute Respiratory Syndrome Coronavirus 2 (SARS-CoV-2), the World Health Organization (WHO) has announced this disease is now an established and ongoing health issue that no longer constitutes a public health emergency of international concern [1]. However, the unceasing circulation and ongoing evolution of SARS-CoV-2 leads to the constant emergence of novel viral lineages that challenge the immune barrier established by previous infections and vaccine boosters, particularly since the emergence of the variant of concern (VOC) Omicron [1]. In the last years, immune evasion has reached a new threshold with the emergence of the Omicron BA.2 descendent lineages XBB and JN.1, which greatly reduced neutralization responses induced by both SARS-CoV-2 wild-type monovalent (wild type) and bivalent (wild type + Omicron BA.5) vaccines [3–8]. The XBB variant resulted from a recombination between two BA.2 lineages (BJ.1 and BM.1.1.1), was first identified in mid-August of 2022 and several XBB descendent lineages (XBB.1*) dominated viral transmissions across the world in 2023 [9–11]. Another substantially mutated BA.2 lineage, the BA.2.86, emerged in August 2023 and carries more than 30 mutations in the Spike (S) protein compared with XBB and BA.2 variants [12]. BA.2.86 rapidly evolved leading to the emergence of several descendant lineages such as JN.1, that emerged in late 2023, which has only one additional mutation in the S protein (L455S) compared with BA.2.86 and it is one of the most immune-evading variants to date [13,14]. The JN.1 family (JN.1*) reached global dominance at the beginning of 2024 and represented more than 90% of the SARS-CoV-2 lineages detected worldwide as of June 2024 [11,15].

In Brazil, the epidemiological scenario regarding the sequential replacements of BA.5 by XBB.1* lineages in early 2023 and of XBB.1* by JN.1* lineages in late 2023, followed the same pattern observed at the global scale [16,17]. Nevertheless, the immediate epidemiological impact of the spread of JN.1* lineages across different Brazilian regions has not been previously explored. In this study we combined data of SARS-CoV-2 sequences with information of severe acute respiratory infection (SARI) cases across different Brazilian states to investigate the potential rise in SARI cases in relation to the timing of XBB.1* replacement by JN.1* lineages. Moreover, we further assessed the cross-reactive neutralizing antibody (NAb) responses against JN.1 in a cohort of convalescent Brazilian patients infected with XBB.1* lineages.

## Materials and Methods

### Sample collection and human cohorts

Nasopharyngeal swabs and serum samples were obtained from 29 individuals (Table S1) selected from a major study that aimed to investigate SARS-CoV-2 dynamics among patients and household contacts in a slum in Rio de Janeiro, Brazil [18,19]. The cohort of volunteers analyzed in the present study is composed of 10 individuals infected with XBB.1* lineages between January and September of 2023, including *i*) six individuals with serum samples collected 25–255 days before and 20-40 days after the infection, and *ii*) four individuals with serum samples collected only 25-30 days after infection. Moreover, serum samples from 19 individuals infected with JN.1* lineages between January and February of 2024, collected 30-70 days after infection, were also included. Nasopharyngeal swabs were used to screen for SARS-CoV-2 by real-time RT-PCR 4Plex SC2/VOC Molecular Kit (Bio-Manguinhos, Rio de Janeiro, Brazil), or TaqPath COVID-19 RT-PCR Kit (Applied Biosystems, Waltham, Massachusetts, EUA), according to the manufacturer’s instructions. Viruses from selected individuals were sequenced by the COVID-19 Fiocruz Genomic Surveillance Network, and all genomic and epidemiological data were uploaded to the EpiCoV™ database (Table S1) [20]. The Brazilian National Committee of Ethics in Research (CONEP) gave ethical approval for this work (protocol number: 30639420.0.0000.5262), and signed written informed consent was obtained from all participants.

### SARS-CoV-2 Brazilian genome sequences

A total of 6,785 SARS-CoV-2 complete genome sequences recovered across different Brazilian states between 1^st^ July 2023 and 31^st^ March 2024 were newly generated by the COVID-19 Fiocruz Genomic Surveillance Network. All samples had real-time RT-PCR cycling thresholds below 30, indicating elevated viral load. SARS-CoV-2 genome sequences were generated using the Illumina COVIDSeq Test kit. Raw data were converted to FASTQ files at Illumina BaseSpace cloud, and consensus sequences were produced with the most up-to-date version of DRAGEN COVID LINEAGE. All genomes were evaluated for mutation calling and quality with the Nextclade 2.14.0 algorithm and were uploaded to the EpiCoV™ database of GISAID (https://gisaid.org/) (see the supplemental material) [21]. Additionally, we downloaded all publicly available Brazilian sequences (*n* = 6,216) collected between 1^st^ July 2023 and 31^st^ March 2024 that were submitted to the EpiCoV™ database until 10 July 2024 with complete collection date/location and lineage assignment. Whole-genome consensus sequences were classified using the “Phylogenetic Assignment of Named Global Outbreak Lineages” (PANGOLIN) software v4.3 (pangolin-data version 1.21) [22].

### SARS-CoV-2 Isolates

Two isolates belonging to the XBB.1.9 (EpiCoV™ accession ID: **EPI_ISL_18250802**) and JN.1.1 (EpiCoV™ accession ID: **EPI_ISL_19225686**) lineages were obtained from nasopharyngeal swabs and used as reference isolates in the Plaque Reduction Neutralization Test (PRNT) assay. Briefly, eligible SARS-CoV-2-positive samples were submitted to virus isolation in Vero E6 cells at a Biosafety level 3 Laboratory, as previously described [23]. When cytopathic effect (CPE) was observed, culture supernatants were collected in working stocks and an aliquot was submitted to real-time RT-PCR followed by whole-genome sequencing for lineage confirmation. Once confirmed, the consensus sequences were deposited at the EpiCoV™ database on GISAID (www.gisaid.org) and each SARS-CoV-2 isolate was titrated by plaque assay.

### PRNT Assay

PRNT was used to determine the serum titers of SARS-CoV-2 neutralizing antibodies, as previously described by Pauvolid-Correa and colleagues [23], with some modifications. Briefly, sera were treated at 56°C for 30 min and used in PRNT in Vero cells (ATCC, CCL 81) maintained in cell culture medium supplemented with fetal bovine serum, sodium bicarbonate, antibiotics/antimycotics, and cultivated at 5% CO_2_ atmosphere and 37°C. Serum samples were tested in duplicates in serial 2-fold dilutions (1:10 to 1:320) for their ability to neutralize 40–60 plaques forming units (PFUs) by each one of the two SARS-CoV-2 reference isolates: XBB.1.9 and JN.1.1. Sera were mixed with the respective SARS-CoV-2 isolates and incubated at 37°C for 1h. Afterward, the culture medium was removed from Vero cell monolayers on 12-well culture plates, which were inoculated with the virus-serum mixtures, incubated at 37°C for 1h, and overlaid with a pre-warmed cell culture medium containing 0.5% ultrapure agarose (Sigma-Aldrich). To ensure reproducibility between experiments and to establish a baseline for neutralization activity, each neutralization experiment included an infection control (no serum) and a reference serum. After 48h of incubation, plates were overlaid with a pre-warmed cell culture medium containing 0.5% ultrapure agarose and neutral red solution (Sigma-Aldrich) to visualize viral plaques, and after 72h, PFUs were counted through a transilluminator. The neutralization titer of a serum is calculated as the reciprocal value of the highest serum dilution that reduces the number of viral plaques by 90% (PRNT_90_). A serum sample is considered reactive to a specific SARS-CoV-2 lineage when the PRNT_90_ neutralization titer reaches a value of at least 10 against that isolate [24].

### Data on hospitalizations for SARI

We extracted data about hospitalizations resulting from SARI attributed explicitly to SARS-CoV-2 (SARI-COVID) in Brazil during the period spanning from July 2023 to March 2024. This information was sourced from the Influenza Surveillance Information System (SIVEP-Gripe) database (https://opendatasus.saude.gov.br/dataset?tags=SRAG). To identify SARI cases, we utilized a set of four criteria which required individuals to exhibit: (i) fever, including self-reported cases; (ii) cough or sore throat; (iii) dyspnea or oxygen saturation levels below 95% or experiencing respiratory discomfort; and (iv) hospitalization. Once an individual meets these criteria and is admitted to a hospital for SARI, their case must be reported and recorded as a distinct entry in the SIVEP-Gripe database. Confirmation of hospitalization for SARI-COVID was contingent on a positive result from the RT-PCR test for SARS-CoV-2. For our analysis, we considered all the records within the SIVEP-Gripe database that adhered to the criteria for defining a hospitalized SARI case. We excluded records related to non-hospitalized deaths from our examination.

### Variant-specific SARI cases

To estimate the number of SARS-CoV-2 infections driven by different variants, we combine the variant frequency data with the number of SARI cases over time (Figure S1). For these analyses, we included a total of 15 states from Northern (Amazonas [AM] and Para [PA]), Northeastern (Alagoas [AL], Ceara [CE], Paraiba [PB], Pernambuco [PE], Piaui [PI], Rio Grande do Norte [RN] and Sergipe [SE]), Central-Western (Goiás [GO] and Federal District [DF]), Southeastern (Minas Gerais [MG], Rio de Janeiro [RJ] and Sao Paulo [SP]) and Southern (Parana [PR], Santa Catarina [SC] and Rio Grande do Sul [RS]) regions that displayed a significant number of SARI cases (n > 100) and clearly defined wave peaks in the study period.

### Data analysis

Graph representations and statistical analysis were performed using GraphPad Prism software v10.2.3 (GraphPad Software, San Diego, CA, USA). The Wilcoxon signed rank test and Mann–Whitney test were used to assess the significant difference between the data sets of PRNT_90_ titers values obtained. A *P* value < 0.05 was considered to be statistically significant.

## Results and Discussion

We analyzed a total of 13,001 Brazilian SARS-CoV-2 genomes sampled across all Brazilian regions and states between 1^st^ July 2023 and 31^st^ March 2024 (Figure 1a and Figure S1). Viral sequences were mostly classified as XBB.1* (63%), JN.1* (31%), or XDR (5%) lineages. XDR is a recombinant clade between XBB.1 and JN.1 descendent lineages which harbors the S protein of JN.1 and display a transmissibility similar to that lineage and was then grouped within the family of viral lineages with a JN.1* S protein [25]. The JN.1* lineages were first detected in Brazil in October 2023 and achieved dominance (>50% of cases) by January 2024. This pattern was quite homogenous across all Brazilian regions and states, except for Ceara, located in the Northeastern region, where the JN.1* lineages already achieved dominance by November 2023 (Figure 1b and Figure S2). This clear pattern of substitution of XBB.1* by JN.1* lineages across all Brazilian localities is consistent with the higher JN.1* transmissibility concerning XBB.1* lineages previously estimated in Brazil and elsewhere [13,25].

**Figure 1.**
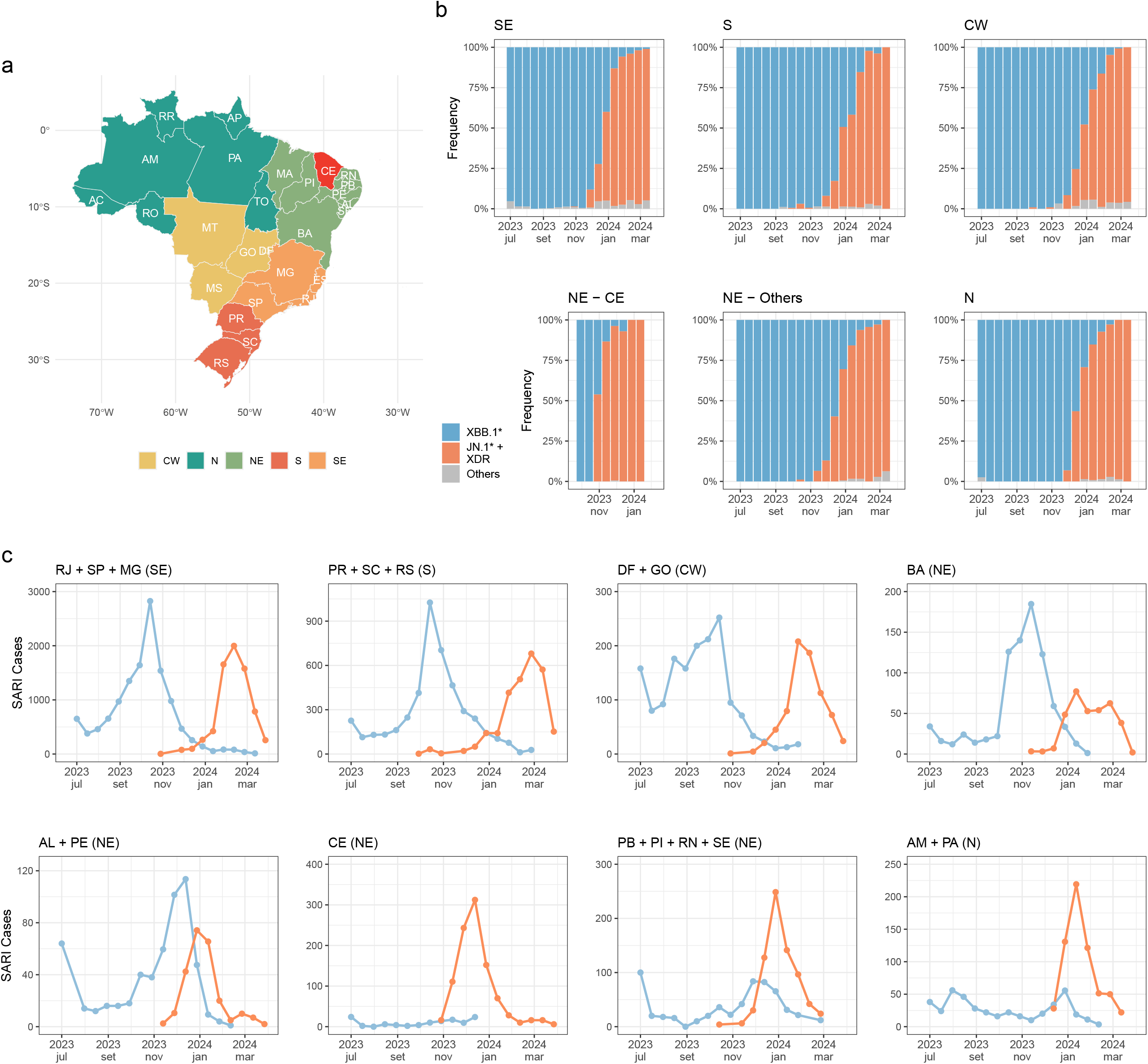
Space-time evolution of major SARS-CoV-2 variants and variant-specific SARI cases in Brazil between July 2023 and March 2024. **a)** The map depicts Brazil’s regions and states, highlighting the Southeastern (SE), Southern (S), Central-Western (CW), Northeastern (NE), and Northern (N) regions in distinct colors as indicated in the legend at the bottom. The state of Ceará (CE) is highlighted in red. Each Brazilian state is labeled with its two-letter abbreviation. **b)** Temporal fluctuations in the relative prevalence of XBB.1* and JN.1*+XDR lineages across Brazilian regions. **c)** Estimated number of SARI cases caused by SARS-CoV-2 XBB.1* and JN.1*+XDR lineages through time across selected Brazilian states from Southeastern (MG, RJ, and SP), Southern (PR, SC, and RS), Northeastern (AL, BA, CE, PB, PE, PI, RN, and SE) and Northern (AM and PA) regions. States from the same region showing a similar pattern of temporal fluctuations of SARS-CoV-2 lineages (**Figure S2**) and SARI cases (**Figure S3**) were grouped for visual clarity. The Brazilian map was generated with the ggplot2 [51], sf [52], and geobr [53] R packages.

To assess the epidemiological consequences of the dissemination of JN.1* lineages across different Brazilian states, we estimated the relative number of variant-specific infections by combining variant frequency data with the number of SARI-COVID cases over time. This analysis revealed four major epidemic patterns (Figure 1c). In some states from Northeastern (CE, PI, PB, RN, and SE) and Northern (AM and PA) regions, the JN.1* lineages started to circulate after a long time interval (> 6 months) of low SARI incidence and viral lineage replacement coincided with a sharp increase of SARI cases that peaked in December 2023-January 2024. In the Northeastern states of AL and PE, viral lineage replacement also coincided with a sharp increase of SARI cases in late 2023, but such a wave was driven by a mixture of both XBB.1* and JN.1* lineages. The XBB variant wave started a couple of weeks earlier and reached a peak somewhat larger than the JN.1 variant wave. The Northeastern state of BA displayed a wave of SARI cases driven by XBB.1* that peaked in November 2023, shortly followed by a wave of smaller size driven by JN.1* that extends from January to February 2024. Finally, states from Central-Western, Southeastern, and Southern regions experienced a wave of SARI cases driven by XBB.1* that peaked around October 2023, a subsequent spread of JN.1* lineages without an immediate upsurge of SARI cases, and a delayed wave of SARI cases driven by JN.1* around February-March 2024.

This pattern observed in the Northeastern/Northern regions resembles that previously described in Brazil during replacements of B.1* lineages by Gamma and of Delta by Omicron (BA.1), although the total number of SARI cases in the JN.1 wave was much lower than in previous waves [26–30]. Meanwhile, the pattern of lineage replacement without an upsurge of SARI cases observed in the other country regions resembles that described during the substitution of VOC Gamma by Delta in Brazil [26,31]. These findings are consistent with the notion that SARS-CoV-2 lineage replacements are primarily driven by the relative transmissibility of viral lineages, while epidemic dynamics also depend on the underlying population immunological landscape and, particularly, the proportion of susceptible hosts [2,32–35]. We hypothesized that the XBB variant wave that occurred in some Brazilian regions around October 2023 boosted the population cross-immunity and generated short-term protection against JN.1 infection, thus preventing an immediate upsurge of SARI cases during lineage turnover.

To test this hypothesis, we investigated the NAb responses induced by XBB.1* infections against the JN.1 variant in 10 Brazilian individuals who were longitudinally followed. As expected, a significant (*P < 0.05*) and large increase in PRNT_90_ titers (29.6-fold) was observed between pre- and post-infection sera against XBB.1 (Figure 2a). We also observed a significant (*P < 0.05*) increase (10.7-fold higher) in the PRNT_90_ titers against JN.1 in post-infection sera compared with the pre-infection samples (Figure 2a). The PRNT_90_ titers against JN.1 were 2.9-fold lower than against XBB.1 after XBB.1* infection, while the PRNT_90_ titers against JN.1 after JN.1* infection were 3.5-fold higher than those obtained after XBB.1* infection (Figure 2b). Despite this reduced neutralizing activity, neutralization of JN.1 was observed in most (90%) XBB-infected individuals analyzed. These results agree with recent studies showing a significant (∼10-fold) increase in NAb responses against the JN.1 variant following XBB.1.5 vaccine booster or XBB.1* infection [36–41]. Those studies also showed that serum titers against JN.1 were around 2□ to 6□fold lower than against XBB.1.5, depending on population immune backgrounds.

**Figure 2.**
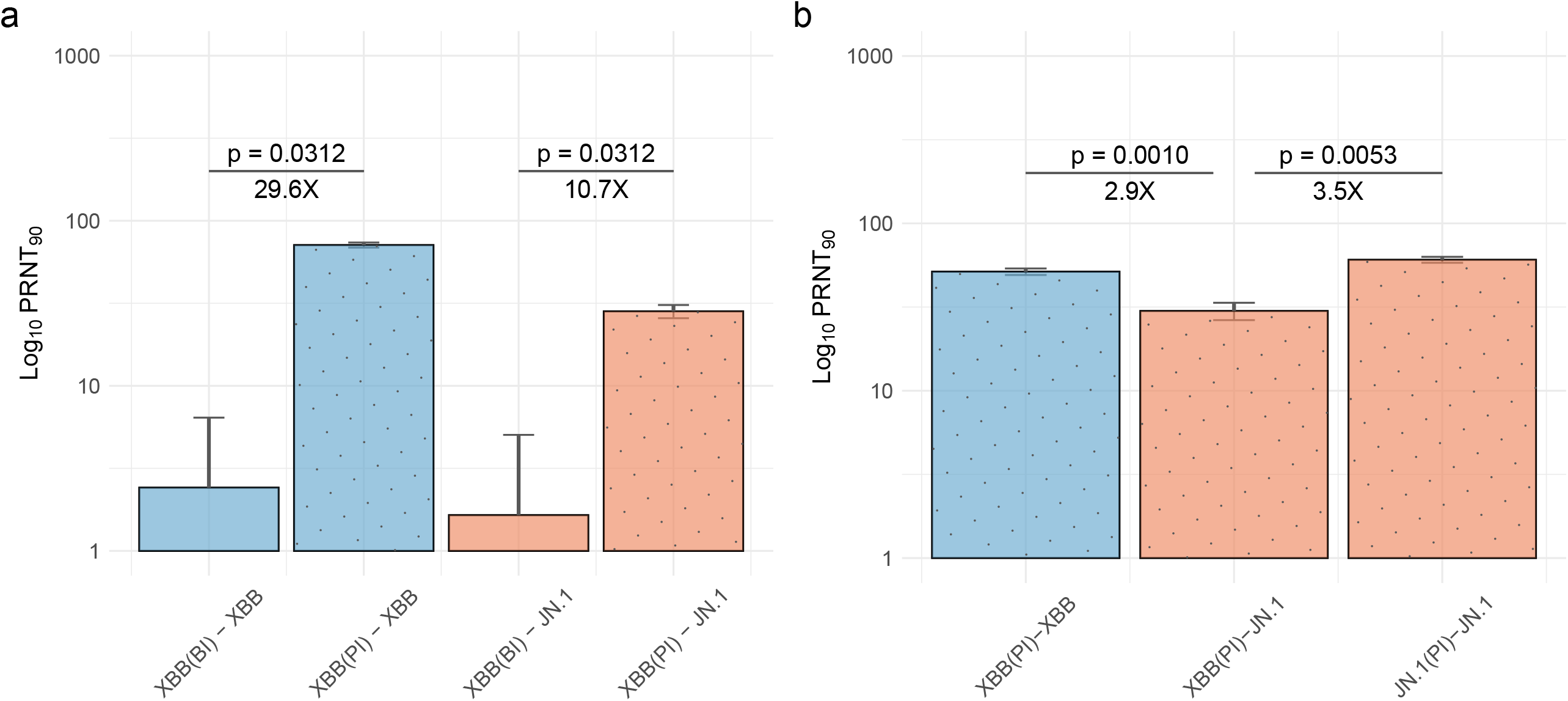
Neutralizing antibodies titers against XBB.1 and JN.1 variants of SARS-CoV-2 in Brazilian individuals who experienced an infection with XBB.1* or JN.1* lineages. **a)** Neutralizing antibodies activity was measured by 90% plaque reduction neutralization titers (PRNT90) against XBB.1 and JN.1 variants in serum samples from a cohort of volunteers (n = 6) that were infected with XBB.1* lineages. Paired serum samples collected before infection - XBB(BI)- and after infection with XBB.1* -XBB(PI)-were tested for each patient. **b)** PRNT90 against XBB.1 and/or JN.1 variants in a set of serum samples from individuals infected with XBB.1* (n = 10) or JN.1* (n = 19) lineages. Samples were collected at 25–255 days before infection (BI) and 20-70 days post-infection (PI). y-axis shows the Log_10_ PRNT_90_ values. Fold-change in the Geometric mean titers (GMT) values and statistical significance are shown. Wilcoxon signed-rank test (A) and Mann–Whitney test (B) were used to calculate the p-values. PRNT_90_ below the limit of detection (< 10) were set to 1. XBB(BI)-XBB = sera obtained BI with XBB.1* and tested against XBB.1. XBB(PI)-XBB = sera PI with XBB.1* and tested against XBB.1. XBB(BI)-JN.1 = sera obtained BI with XBB.1* and tested against JN.1. XBB(PI)-JN.1 = sera obtained PI with XBB.1* and tested against JN.1, JN.1(PI)-JN.1 = sera obtained PI with JN.1* and tested against JN.1. The bars are color-coded based on the isolate used in each category, with blue representing XBB* isolates and orange for JN.1 isolates. PI sera bars are depicted with a dotted pattern.

The epidemic pattern of SARI cases in the Southeastern/Southern/Central-Western regions reveals a minimum time interval of 4-5 months between the XBB and JN.1 wave peaks (Figure 1c). This is consistent with the notion that cross-reactive immunity induced by XBB.1* infections wanes over time, fueling a JN.1 wave of SARI cases ∼4-5 months later in those regions [42,43]. Interestingly, among the 19 individuals from the Southeastern state of RJ infected with the JN.1 variant analyzed in our study, seven had a documented previous SARS-CoV-2 infection (Table S1). This includes three individuals who were infected during the period of XBB dominance between March and September 2023. The time interval between previous XBB infections and JN.1 reinfections ranges between 5.3 and 10.8 months. These findings corroborate that individuals previously infected with XBB.1* lineages could be reinfected with JN.1* lineages and further support a minimum time interval of about five months between successive infections by those Omicron lineages in the Brazilian population immune landscape. Despite the small number of subjects analyzed, our findings support that XBB.1* infections produced substantial cross-reactive NAb responses against the JN.1 variant in the Brazilian immune background. The precise level of NAb required for protection against JN.1 infection is difficult to estimate, but some evidence supports that such cross-reactive responses induced by XBB may be protective. A recent report showed that immunity provided by natural infection against reinfection with JN.1 was strong (83% effectiveness) among those who were infected within the last six months with variants such as XBB.1*[44]. Other studies also showed that XBB.1.5 monovalent mRNA vaccine booster conferred some protection against symptomatic infections and hospitalizations caused by the JN.1 variant [45–49]. In Brazil, the XBB.1.5 monovalent mRNA vaccine was first introduced by the end of May 2024 [50]. Our findings, along with previous studies, emphasize the importance of boosters with updated XBB.1.5 monovalent vaccines to enhance immune responses and reduce onward transmission of the JN.1 variant currently prevalent in Brazil, particularly in priority groups of the population.

Our study has some limitations. First, we estimated the relative number of variant-specific infections assuming that SARS-CoV-2 genomic surveillance was random and truly reflected the variant frequency within states and that the infection-hospitalization ratio (SARI cases/total cases) was the same for different Omicron variants. Second, due to the small sample size of XBB-infected individuals, we could not assess the potential impact of age or immune background (history of infections and vaccinations) on neutralization efficiency. XBB-infected individuals’ ages range between 40 and 89 (Table S1), pointing out that cross-reactive responses were not restricted to young adults. Moreover, all XBB-infected individuals included in our study were fully vaccinated (monovalent wild-type vaccines) and a significant fraction (70%) also have a documented previous infection (Table S1), thus indicating that hybrid immunity was an important component of the immune background of the studied population. Third, serum samples were collected within two months post XBB.1* infection and we do not have information about cross-reactive neutralization efficiency against JN.1 after longer periods since infection.

## Conclusions

The JN.1* lineages consistently replaced the XBB.1* lineages circulating in all Brazilian states since late 2023 but with variable epidemiological impact across regions. In those Brazilian states that experienced a XBB variant wave in the second half of 2023, shortly before the introduction of JN.1, lineage replacement occurred without a concurrent rise in the number of SARI cases. By contrast, Brazilian states without a recent XBB variant wave in the second half of 2023 experienced an upsurge of SARI cases concurrent with the expansion of the JN.1 variant. Recent infections with XBB.1* lineages induce a considerable production of cross-reactive NAb that are effective against JN.1 in the Brazilian population’s immune background. Together, these results support the notion that the XBB variant wave observed in the second half of 2023 in some Brazilian states probably acted as a natural booster of the population immunity, providing short-term protection against JN.1 infections and delaying the rise of SARI cases in those country regions.

## Data Availability

This study’s conclusions derive from examining 13,001 SARS-CoV-2 genomes from Brazil, made publicly accessible via the EpiCoV™ database from GISAID. These genomes were collected after July 1^st^, 2023, and submissions were recorded until March 31^st^, 2024. The data can be accessed at https://doi.org/10.55876/gis8.240719ev.

## Acknowledgments

We would like to thank the Biosafety Laboratory Level 3 platform (BSL3) at Pavilhão *Helio Peggy Pereira*, Oswaldo Cruz Institute, FIOCRUZ, Rio de Janeiro, Brazil.

