## Supplementary material for "SARS-CoV-2 Omicron XBB infections boost cross-variant neutralizing antibodies, potentially explaining the observed delay of the JN.1 wave in some Brazilian regions"

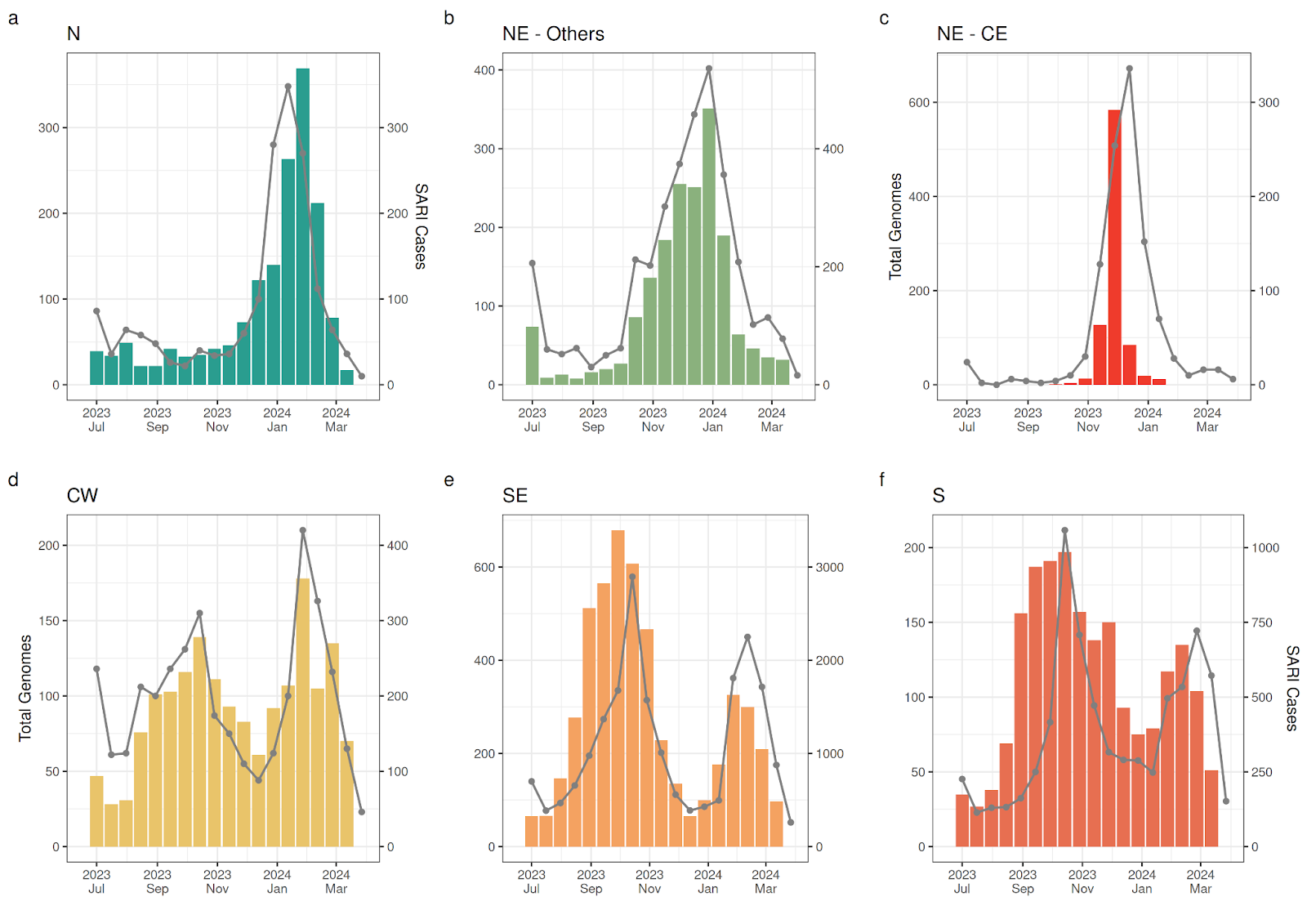

**Figure S1. SARS-CoV-2 genomic sampling and severe acute respiratory infection (SARI) cases in Brazil from July 2023 to March 2024.** This figure presents the biweekly number of available SARS-CoV-2 genome samples from Brazil (n_TOTAL_ = 13,001, left y-axis and colored bars) alongside SARI cases (n_TOTAL_ = 13,001, right y-axis and dotted gray lines) recorded during the study period. The data is stratified by Brazil’s geographic regions (**a-f**). The northeastern state of Ceará (**c**) is highlighted separately due to its substantial contribution to the region's genomic dataset in late 2023, comprising 55% of the Northeastern samples in December 2023.

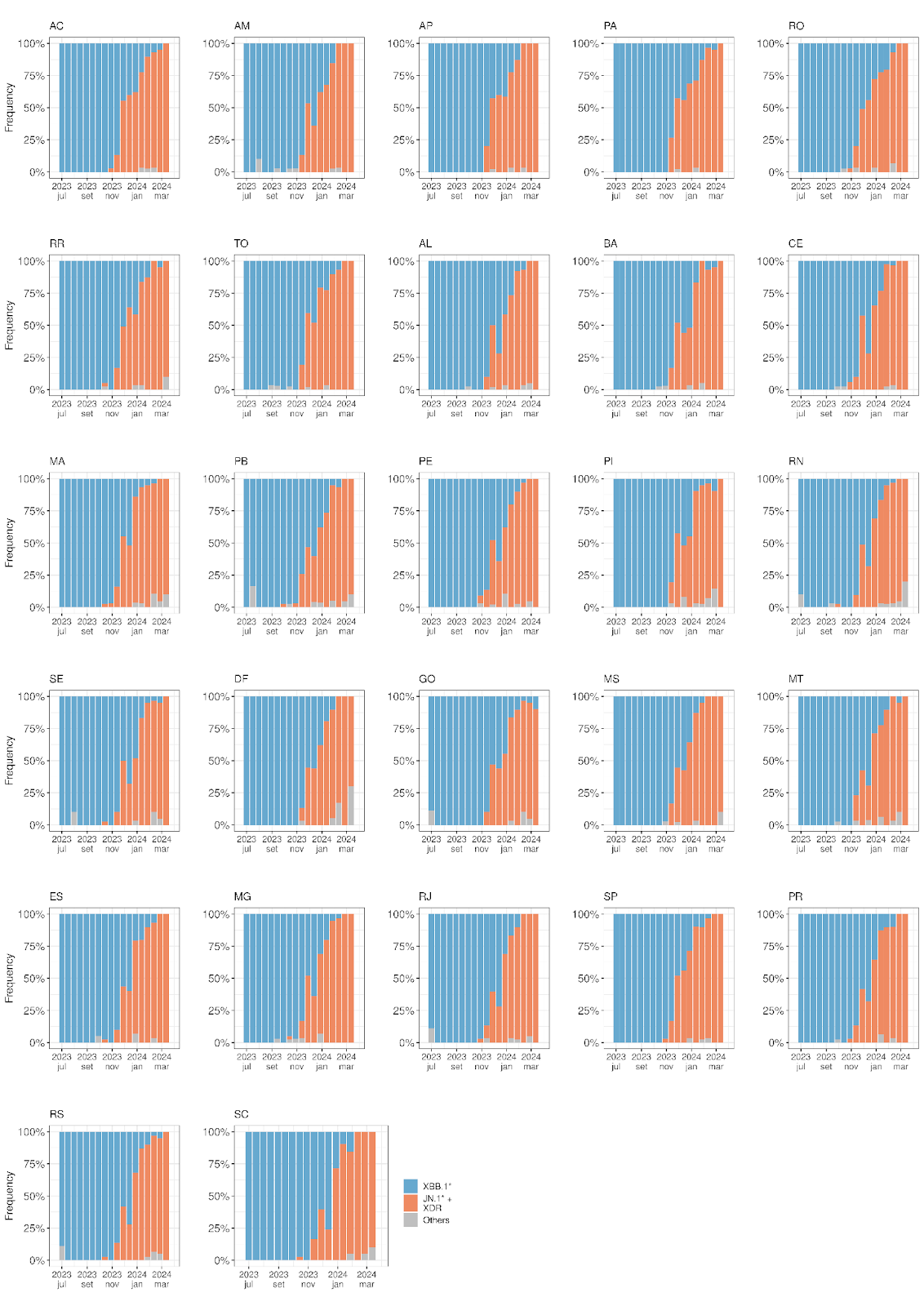

**Figure S2. SARS-CoV-2 genomic profile in Brazil from July 2023 to March 2024.** The figure presents the biweekly Pango lineages composition of SARS-CoV-2 genomes sampled across Brazil's 27 federative units, including all 26 states and the Federal District (DF). The figure follows the color scheme on the last panel's right side.

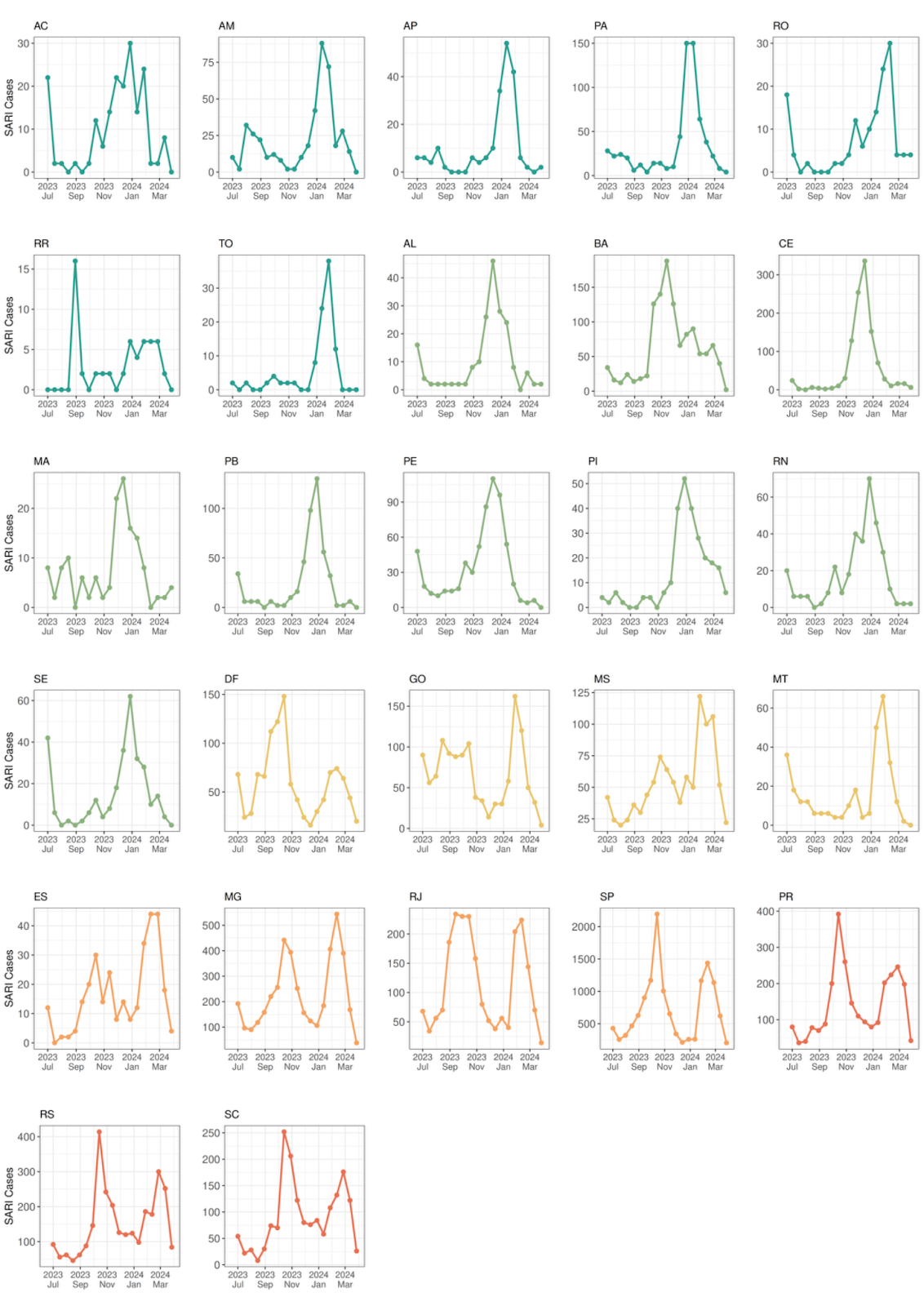

**Figure S3. SARI-COVID cases in Brazil from July 2023 to March 2024.** The figure illustrates the weekly incidence of SARI (severe acute respiratory infection) cases associated with SARS-CoV-2 infection reported across Brazil's 27 federative units, encompassing all 26 states and the Federal District (DF). States from Northern (dark green), Northeastern (light green), Centra-Western (yellow), Southeastern (orange), and Southern (red) regions are represented by different colors.

**Table S1. Genomic and epidemiological data of individuals analyzed in this study.**

| **Age**  **ranges** | **Sex** | **Previous Vaccination date** | **Previous infection date** | **Lineage^a^** | **XBB/JN.1 Infection date** | **Lineage^b^** | **Access ID** | **Sera collection date** | **Days**  **from XBB infection** |
| --- | --- | --- | --- | --- | --- | --- | --- | --- | --- |
| 80-89 | female | Jun-22  (Pfizer mono**^c^**) | Jan-22 | unknown | Jan-23 | XBB.1.18 | EPI_ISL_17471750 | 1st (Nov-22)  2nd (Fev-23) | 1st: -66  2nd: +20 |
| 70-79 | male | Apr-22 (Janssen**^d^**) | Nov-22 | BA.1.15 | Feb-23 | XBB.1.5.15 | EPI_ISL_18453820 | 1st (Dec-22)  2nd (Mar-23) | 1st: -70  2nd: +37 |
| 50-59 | female | Jun-22 (Janssen) | Jan-22 | unknown | Feb-23 | XBB.1.5.15 | EPI_ISL_17471744 | 1st (Jan-23)  2nd (Mar-23) | 1st: -38  2nd: +32 |
| 40-49 | male | Nov-22  (Pfizer mono) | unknown | n.a. | Mar-23 | XBB.1.5 | EPI_ISL_18453821 | Mar-23 | +27 |
| 40-49 | female | Jul-22 (AstraZeneca**^e^**) | Jan-22 | unknown | Mar-23 | XBB.1.15.1 | EPI_ISL_18453823 | 1st (Jun-22)  2nd (Mar-23) | 1st: -252  2nd: +23 |
| 40-49 | female | Nov-22  (Pfizer mono) | Mar-21 | unknown | Mar-23 | XBB.1.5.17 | EPI_ISL_18453822 | 1st (Feb-23)  2nd (Apr-23) | 1st: -27  2nd: +35 |
| 40-49 | female | Jun-22  (Pfizer mono) | Mar-21 | unknown | Mar-23 | XBB.1.5 | EPI_ISL_18453825 | Apr-23 | +27 |
| 60-69 | female | Mar-23  (Pfizer bi**^f^**) | Nov-20 | unknown | May-23 | XBB.1.5.70 | EPI_ISL_18453828 | 1st (Apr-23)  2nd (Jun-23) | 1st:-31  2nd: +32 |
| 40-49 | male | Mar-23  (Pfizer bi) | unknown | n.a. | Sep-23 | XBB.1.16.6 | EPI_ISL_18345911 | Oct-23 | +28 |
| 40-49 | male | Dec-21  (Pfizer mono) | unknown | n.a. | Sep-23 | XBB.1.16.6 | EPI_ISL_18415723 | Oct-23 | +27 |
| 60-69 | female | Mar-23  (Pfizer bi) | May-23 | XBB.1.5.70 | Jan-24 | JN.1 | EPI_ISL_18934171 | Mar-24 | +42 |
| 60-69 | female | May-22 (Janssen) | no infection | n.a. | Jan-24 | JN.1.16 | EPI_ISL_18934174 | Mar-24 | +40 |
| 70-79 | female | Mar-23  (Pfizer bi) | Jun-20 | unknown | Jan-24 | JN.1 | EPI_ISL_18934178 | Mar-24 | +41 |
| 40-49 | female | Apr-23  (Pfizer bi) | Mar-23 | XBB.1.5.17 | Jan-24 | JN.1 | EPI_ISL_18934179 | Apr-24 | +66 |
| 40-49 | female | Mar-23  (Pfizer bi) | unknown | n.a. | Jan-24 | JN.1 | EPI_ISL_18952125 | Mar-24 | +35 |
| 20-29 | male | Jun-23  (Pfizer bi) | unknown | n.a. | Feb-24 | JN.1 | EPI_ISL_18952126 | Mar-24 | +39 |
| 40-49 | female | Jul-23  (Pfizer bi) | unknown | n.a. | Feb-24 | JN.1 | EPI_ISL_18952130 | Mar-24 | +41 |
| 40-49 | male | Mar-23  (Pfizer bi) | unknown | n.a. | Feb-24 | JN.1.4 | EPI_ISL_18952127 | Mar-24 | +49 |
| 20-29 | female | Mar-23  (Pfizer bi) | no infection | n.a. | Feb-24 | JN.1.4 | EPI_ISL_18952129 | Mar-24 | +35 |
| 40-49 | female | Oct-22  (Pfizer mono) | Nov-22 | unknown | Feb-24 | JN.1 | EPI_ISL_18952134 | Mar-24 | +43 |
| 60-69 | female | Mar-23  (Pfizer bi) | unknown | n.a. | Feb-24 | JN.1.4 | EPI_ISL_18971502 | Apr-24 | +49 |
| 50-59 | female | Mar-23  (Pfizer bi) | unknown | n.a. | Feb-24 | JN.1.7 | EPI_ISL_18971463 | Mar-24 | +36 |
| 20-29 | male | Nov-21  (Pfizer mono) | unknown | n.a. | Feb-24 | JN.1 | EPI_ISL_18971464 | Apr-24 | +41 |
| 30-39 | male | Aug-22  (Pfizer mono) | no infection | n.a. | Feb-24 | JN.1 | EPI_ISL_18971496 | Apr-24 | +42 |
| 30-39 | female | May-23  (Pfizer bi) | Sep-23 | unknown | Feb-24 | JN.1 | EPI_ISL_18971475 | Apr-24 | +38 |
| 40-49 | female | Mar-23  (Pfizer bi) | May-21 | unknown | Feb-24 | JN.1 | EPI_ISL_18971462 | Apr-24 | +35 |
| 1-9 | female | Feb-23  (Pfizer mono) | no infection | n.a. | Feb-24 | JN.1 | EPI_ISL_18971514 | Apr-24 | +35 |
| 70-79 | female | Mar-23  (Pfizer bi) | no infection | n.a. | Feb-24 | JN.1 | EPI_ISL_18971481 | Apr-24 | +35 |
| 40-49 | female | May-23  (Pfizer bi) | Nov-22 | BF.15 | Feb-24 | JN.1 | EPI_ISL_18971506 | Apr-24 | +32 |

**^a^**Lineage of previous infection.

**^b^**Specific lineage of XBB/JN.1 Infection.

**^c^***Pfizer mono*, refers to the Pfizer-BioNTech COVID-19 Vaccine – Comirnaty, BNT162b2.

**^d^***Janssen*, refers to the Janssen COVID-19 Vaccine, Ad26.COV2.S.

**^e^***AstraZeneca*, refers to the AZD1222/COVISHIELD COVID-19 Vaccine.

**^f^***Pfizer bi*, refers to the Pfizer-BioNTech COVID-19 Vaccine, Bivalent, BNT162b2 (Original/Omicron BA.4/BA.5 or Original/Omicron BA.1).
